# Multidimensional analysis of the clinical spectrum and symptom burden of unexplained myofascial pain

**DOI:** 10.64898/2026.03.27.26349456

**Authors:** Siddhartha Sikdar, Secili DeStefano, María José Guzmán Pavón, Yu-Lin Hsu, Seiyon Lee, John Srbely, Jay Shah, William Rosenberger, Samuel Acuña, Yonathan Assefa, Matin Jahani Jirsaraei, Antonio Stecco, Lynn H. Gerber

**Author notes:** Corresponding Author: Siddhartha Sikdar, PhD, Department of Bioengineering, Center for Advancing Systems Science and Bioengineering Innovation, 4400 University Dr. MS 1J7, Fairfax, VA 22030, USA. Abstracts related to this study were presented at the Padova Myofascial Pain Symposium in July, 2025 and will be presented at the AAPMR annual assembly to be held in Oct, 2025. **Funding:** This analysis was part of a study funded by the NIH through grant 1R61AT012286. Clinical Trials Registration: NCT#: NCT06060925.

## Abstract

**Objective:** Myofascial pain (MP) is a leading cause of disability globally. Pain quality and severity vary widely for people with MP, making it difficult to accurately assess the spectrum of symptoms and develop appropriate treatments. We assessed potential contributors to variability in the clinical spectrum of unexplained neck/shoulder pain and associated myofascial component(s).

**Design:** Prospective cross-sectional study of adults reporting neck/shoulder pain and pain-free individuals.

**Outcomes Measures:** Pain intensity and interference (PEG); Symptom burden measured using patient-reported outcomes and objective measures: pain catastrophizing (PCS); PROMIS physical function (PF); sleep disturbance; anxiety (GAD-2); depression (PHQ-2); hypermobility (Beighton/Brighton); Objective measures in the medial upper trapezius: pressure pain threshold (PPT) and quantitative sensory testing (QST).

**Results:** Of the 96 adults recruited for the study, 82 had complete records (age 32.2 +/−13.1 years, 57% women). On physical exam, 23 were assessed to be in an *active* group (those with spontaneous MP without provocation), 38 in a *latent* group (those with MP upon provocation), and 21 in a *normal* group (no MP in neck and shoulder). The symptom burden explained 75% of the variance in PEG in the overall sample, 85% in the *active* group and 92% in the *normal* group. PF and PCS are key predictors of PEG. Network analysis identified unique symptom clusters in the *active* and *latent* groups.

**Conclusions:** The symptom burden explains the variability in the clinical spectrum of pain intensity and interference in unexplained neck/shoulder MP. Network analysis can further improve clinical risk stratification. These findings represent a step towards an eventual goal of developing multidisciplinary clinical guidance for managing the whole patient, rather than the current emphasis on regional pain contributors in MP.

## Introduction

Chronic musculoskeletal pain (cMSKP) is a leading cause of disability worldwide. Most patients lack a clearly identifiable underlying cause of their symptoms^3^. The traditional approach to studying cMSKP focuses on regional anatomy (e.g., low back, neck, shoulder, knee pain). Myofascial tissues are commonly involved in cMSKP, and are often a treatment target^4–8^. There is no definitive test to diagnose myofascial pain (MP), which is characterized by a motor abnormality (physical findings within the muscle) and a sensory abnormality (tenderness and referred pain)^7,8^. Diagnosis is commonly based on clinical assessments, including pain regionality, presence of palpable, symptomatic myofascial trigger points (MTrPs) in affected tissues, history, and physical examination^9^. Strong digital pressure on symptomatic MTrPs often exacerbates the patient’s spontaneous pain complaint and mimics the patient’s familiar pain experience. This approach has low to moderate reproducibility and reliability^10,11^. The symptom burden exhibits large variability between individuals and over time, and the presence or absence of local MTrPs does not fully capture the complexity of the patient’s lived daily experience.

It remains unclear whether MP is a distinct regional syndrome, or part of the clinical spectrum (and component) of non-specific cMSKP^12,13^. Evidence suggests that the clinical manifestations, lived experience and pain-related functional limitations of cMSKP (and chronic MP) are strongly influenced by physiological, functional and psychosocial determinants^14,15^. The prevailing focus on regional contributing factors fails to address these complex biopsychosocial interconnections, and has led to overly simplified treatment approaches with limited effectiveness for patient outcomes^16^. Inconsistent nomenclature^17^ further adds to the challenges for clinicians, patients and researchers. A growing body of research points to the need for a paradigm shift towards considering chronic pain as an emergent property of a complex system with central and peripheral components^18–24^.

Network analysis is an emerging approach to studying complex systems by identifying system components (nodes) and their relationships (edges)^25^. However, to our knowledge, no previous study has 1) identified whether multidomain biopsychosocial determinants can explain the clinical spectrum of MP; nor 2) whether the network dependencies among these outcomes differs between the clinically defined phenotypes of MP (active, latent) versus normal.

To address this gap, our primary objective was to investigate whether the symptom burden, measured using a set of patient reported outcomes from a standardized chronic pain common data elements inventory^26^, along with validated measures of hypermobility and sensitization, can explain the variability in the clinical spectrum of pain intensity and interference in MP. Our secondary objective was to determine whether the network dependencies differ between *active* MP, *latent* MP and *normal* clinical phenotypes.

## Materials and methods

### Study design

This cross-sectional study was conducted at George Mason University as part of a larger study to develop biomarkers for MP (clinicaltrials.gov NCT#: NCT06060925). For differentiating between two clinically-relevant subgroups with an area under the receiver operating curve of 0.75, we determined that at least 19 subjects in each subgroup are needed to obtain a power of 0.8 and significance of 0.05. Participants were recruited from the general community, and a variety of different healthcare private practices in the greater Virginia/Washington DC/Baltimore area, from January, 2023 to December, 2024. All study procedures were approved by our Institutional Review Board and informed consent was obtained from all participants.

### Participant assessment

Figure 1 shows the flowchart for participant recruitment and assessment.

**Figure 1.**
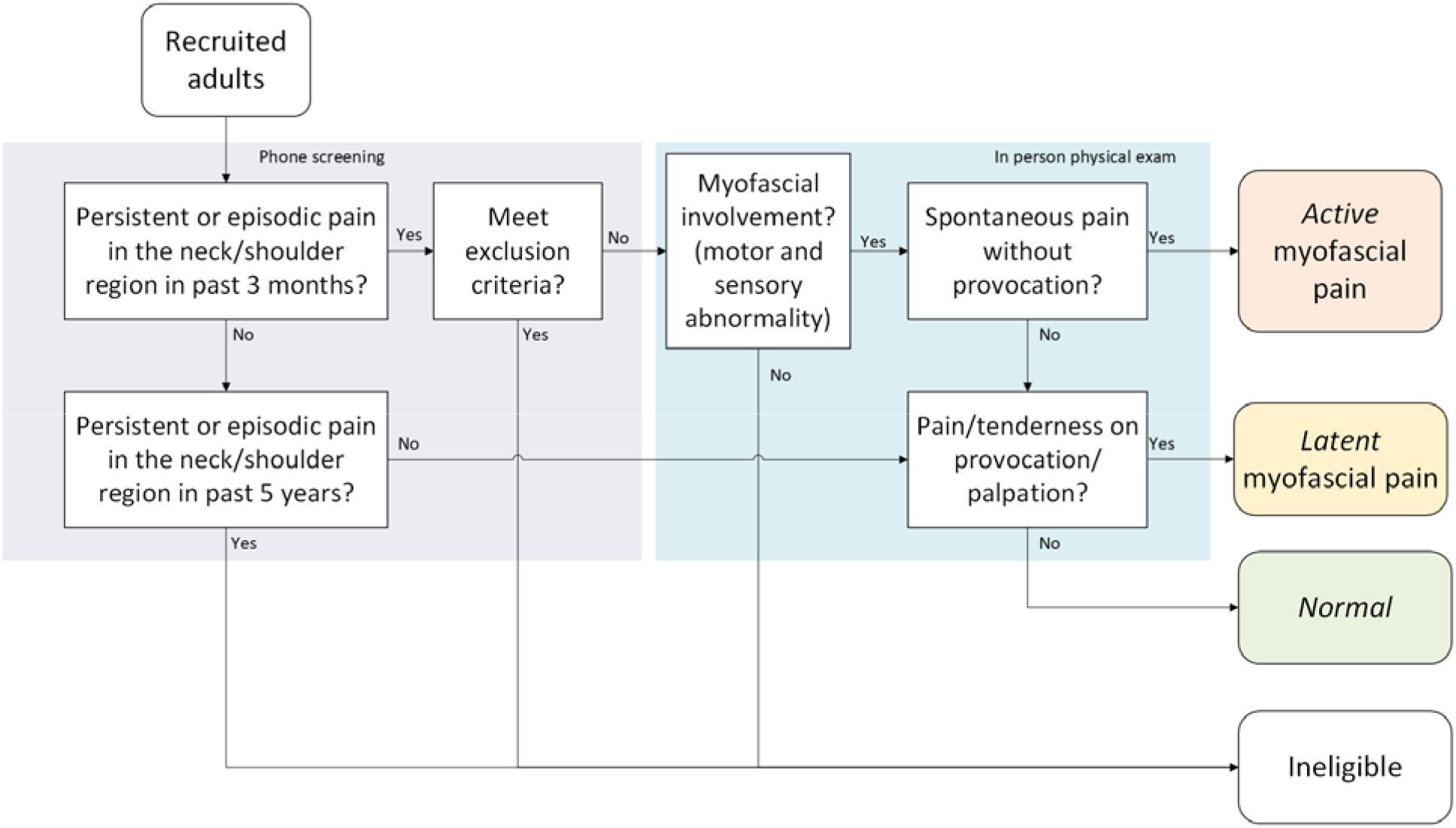
Patient assessment flowchart

#### Phone screening

We recruited adult participants (≥18 years) with persistent or episodic pain in the neck/shoulder region for at minimum the past 3 months, as well as individuals without pain (no self-reported persistent or episodic neck or shoulder pain for the past 5 years). Participants who reported the following during phone screening were excluded: 1) previous diagnosis of fibromyalgia, chronic fatigue syndrome or chronic Lyme disease; 2) previous diagnosis of cervical radiculopathy, neuropathy or neuritis; 3) history of recent head, neck or shoulder girdle surgery; 4) new medication or change in medication in past 6 months; 5) history of shoulder or cervical fracture.

#### Physical examination

For participants meeting eligibility criteria, an in-person comprehensive history and musculoskeletal physical exam was performed by experienced clinicians (two board-certified physical medicine and rehabilitation physicians and two licensed physical therapists). The standardized physical examination^9^ included identification of active and latent MTrPs in the neck and shoulder region following Travell and Simon’s criteria^27^. Clinicians noted findings including the presence of MTrPs, “knots”, taut bands, asymmetry of palpation compared to contralateral side, the presence of a motor abnormality (physical finding in the muscle tissue) and a sensory abnormality (tenderness and referred symptomatic pain)^7,8^. Based on this in-person physical examination, subjects with neck/shoulder pain but without myofascial involvement were excluded (Figure 1).

#### Classification of Myofascial Pain Group

Based on eligibility screening and physical examination, subjects were classified as either active (spontaneous persistent or episodic myofascial pain without provocation in the neck/shoulder region ≥3 months), *latent* (myofascial pain in the neck/shoulder region that is reproduced only upon provocation/palpation), or *normal* (clinically asymptomatic; no self-reported chronic pain in past 5 years; no palpable findings that reproduce pain in the neck/shoulder region; may have asymptomatic palpable findings). Determination of MP was therefore made at the whole-person level, i.e., *active, latent* or *normal* subject, rather than at the regional level, i.e., presence of an active or latent MTrP. This was done to allow analysis using other whole-person measures.

### Outcome Measures

Details on outcome measures are provided in the Appendix.

#### Patient reported outcome measures

We utilized the following standardized adult chronic pain NIH HEAL CDE^26^: Pain, Enjoyment of Life and General activity score (PEG)^28^; Pain Catastrophizing Scale (PCS)^29^; PROMIS Physical Function^30^; PROMIS Sleep Disturbance Scale^31^; Generalized Anxiety Disorder Scale 2-item scale (GAD-2)^32^; Patient Health Questionnaire 2 (PHQ-2)^32^.

#### Modified Beighton-Brighton score^33^

Beighton score^34^ modified with Brighton^35^ to assess for hypermobility syndrome.

#### Sensitization

Pressure pain threshold (PPT)^36,37^ and quantitative sensory testing (QST) for windup^38^.

### Statistical Analysis

Regression analysis was designed to evaluate whether the symptom burden, measured using the HEAL CDE^26^, along with validated measures of hypermobility and sensitization, can explain the variability in the clinical spectrum PEG in MP. Network analysis was used to investigate partial correlations among the different measures.

Only subjects with complete data records for all selected variables were included in this analysis. Summary statistics are presented in Table 1 and in Figure 2. Linear regression analyses were conducted with PEG as the dependent variable. Predictors were GAD-2, hypermobility, PCS, PHQ-2, physical function, sleep disturbance, PPT, and QST. The model was adjusted for sex at birth (male/female), age (considered continuous), and pain groups (*active, latent and normal*). Model aptness was verified using residual plots, and multicollinearity was assessed by analysis of variance inflation factors. Subgroup analyses were performed as warranted.

**Table 1.** Sample characteristics (n=82). Summary statistics report means and standard deviations for continuous variables and percentages for categorical variables.

|  | *Active* | *Latent* | *Normal* | Total |
| --- | --- | --- | --- | --- |
| **Age<sup>¶¶</sup>** | 35.04 (12.17) | 30.45 (12.38) | 32.38 (15.19) | 32.23 (13.08) |
| **Sex at Birth** |  |  |  |  |
| Female | 17 (73.91%) | 18 (47.37%) | 12 (57.14%) | 47 (57.32%) |
| Male | 6 (26.09%) | 20 (52.63%) | 9 (42.86%) | 35 (42.68%) |
| **Race (categories are not exclusive)** |  |  |  |  |
| American Indian/Alaskan Native | 0 (0.0%) | 0 (0.0%) | 0 (0.0%) | 0 (0.0%) |
| Asian | 9 (39.13%) | 16 (42.11%) | 7 (33.33%) | 32 (39.02%) |
| Black or African American | 1 (4.35%) | 2 (5.26%) | 5 (23.81%) | 8 (9.76%) |
| Native Hawaiian or Other Pacific<br>Islander | 0 (0.0%) | 0 (0.0%) |  | 0 (0.0%) |
| White | 13 (56.52%) | 21 (55.26%) | 9 (42.86%) | 43 (52.44%) |
| Not Reported | 0 (0.0%) | 1 (2.63%) | 1 (4.76%) | 2 (2.44%) |
| **Currently experiencing neck/shoulder pain<sup>#</sup>** |  |  |  |  |
| Bilateral pain (%) | 19 (82.6%) | 11 (28.9%) | 2 (9.5%) | 32 (39%) |
| Unilateral pain (%) | 3 (13.1%) | 7 (18.4%) | 2 (9.5%) | 12 (14.6%) |
| No pain (%) | 1 (4.3%) | 18 (47.4%) | 17 (81%) | 36 (43.9%) |
| **Current neck/shoulder pain location<sup>#</sup>** |  |  |  |  |
| Neck and shoulder both (%) | 18 (78.3%) | 7 (18.4%) | 3 (14.3%) | 28 (34.1%) |
| Neck only (%) | 3 (13.0%) | 7 (18.4%) | 1 (4.8%) | 11 (13.4%) |
| Shoulder only (%) | 0 | 8 (21.1%) | 1 (4.8%) | 9 (11.0%) |
| Neither (%) | 0 | 13 (34.2%) | 16 (76.2%) | 29 (35.4%) |
| Not reported (%) | 2 (8.7%) | 3 (7.9%) | 0 | 5 (6.1%) |
| **Physical findings on palpation<sup>§</sup>** |  |  |  |  |
| Bilateral findings (%) | 23 (100%) | 34 (89.5%) | 8 (38.1%) | 65 (79.3%) |
| Unilateral findings (%) | 0 (0%) | 4 (10.5%) | 5 (23.8%) | 9 (11%) |
| No palpable findings (%) | 0 (0%) | 0 | 8 (38.1%) | 8 (9.7%) |
| **Myofascial pain type<sup>¶</sup>** |  |  |  |  |
| Active myofascial pain | 23 | 0 | 0 | 23 (28.1%) |
| Latent myofascial pain | 0 | 38 | 0 | 38 (46.3%) |
| Normal | 0 | 0 | 21 | 21 (25.6%) |
| **Clinical Measures** |  |  |  |  |
| GAD-2<sup>1</sup> | 2.00 (1.86) | 1.24 (1.26) | 0.81 (1.54) | 1.34 (1.57) |
| Hypermobility<sup>2</sup> | 6.87 (4.49) | 4.39 (2.68) | 4.24 (2.76) | 5.05 (3.45) |
| PCS<sup>3</sup> | 7.35 (5.05) | 4.76 (4.81) | 6.52 (5.90) | 5.94 (5.23) |
| PEG<sup>4</sup> | 4.74 (2.18) | 1.11 (1.56) | 0.97 (1.48) | 2.09 (2.39) |
| PHQ-2<sup>5</sup> | 1.48 (1.50) | 0.82 (1.06) | 0.76 (1.34) | 0.99 (1.29) |
| Physical Function<sup>6</sup> | 24.52 (4.49) | 29.37 (1.24) | 28.86 (3.42) | 27.88 (3.68) |
| PPT<sup>7</sup> | 6.33 (5.01) | 6.91 (3.07) | 7.62 (3.97) | 6.93 (3.90) |
| QST<sup>8</sup> | 459.76 (516.54) | 235.99 (273.59) | 213.00 (228.10) | 292.87 (361.37) |
| Sleep Disturbance<sup>9</sup> | 15.22 (4.82) | 12.82 (4.74) | 10.52 (2.99) | 12.90 (4.67) |

<sup>#</sup>Based on participant self report; Note that some subjects clinically classified as *normal* based on the criteria described in the Methods section, reported current neck and shoulder pain.
<sup>§</sup>Based on physical exam by a trained clinician. Physical findings include presence of MTrPs, knots, taut bands, asymmetry compared to contralateral side, etc.
<sup>¶</sup>Myofascial involvement was defined as presence of both a motor abnormality (physical findings on palpation) and a sensory abnormality (tenderness and referred pain).
<sup>1</sup>Scores ≥3 considered indicative of clinical anxiety.
<sup>2</sup>Score categories: severe (11 or more), high (8-10), moderate (4-7), low (2-3), or none (0-1).
<sup>3</sup>Scores ≥30 indicative of clinically-relevant catastrophizing.
<sup>5</sup>Scores ≥3 are typically considered clinically-relevant, although a threshold of 2 has also been recommended.
<sup>6</sup>Score = 30 indicates no limitation in physical function. Lower scores are worse.
<sup>7</sup>Minimum pressure (in lb/sq inch) needed to evoke discomfort or pain. Lower scores are worse.
<sup>8</sup>Score computed as sum square of numerical pain ratings for 16 consecutive pinpricks. Higher scores are worse.
<sup>9</sup>Higher scores are worse.

**Figure 2.**
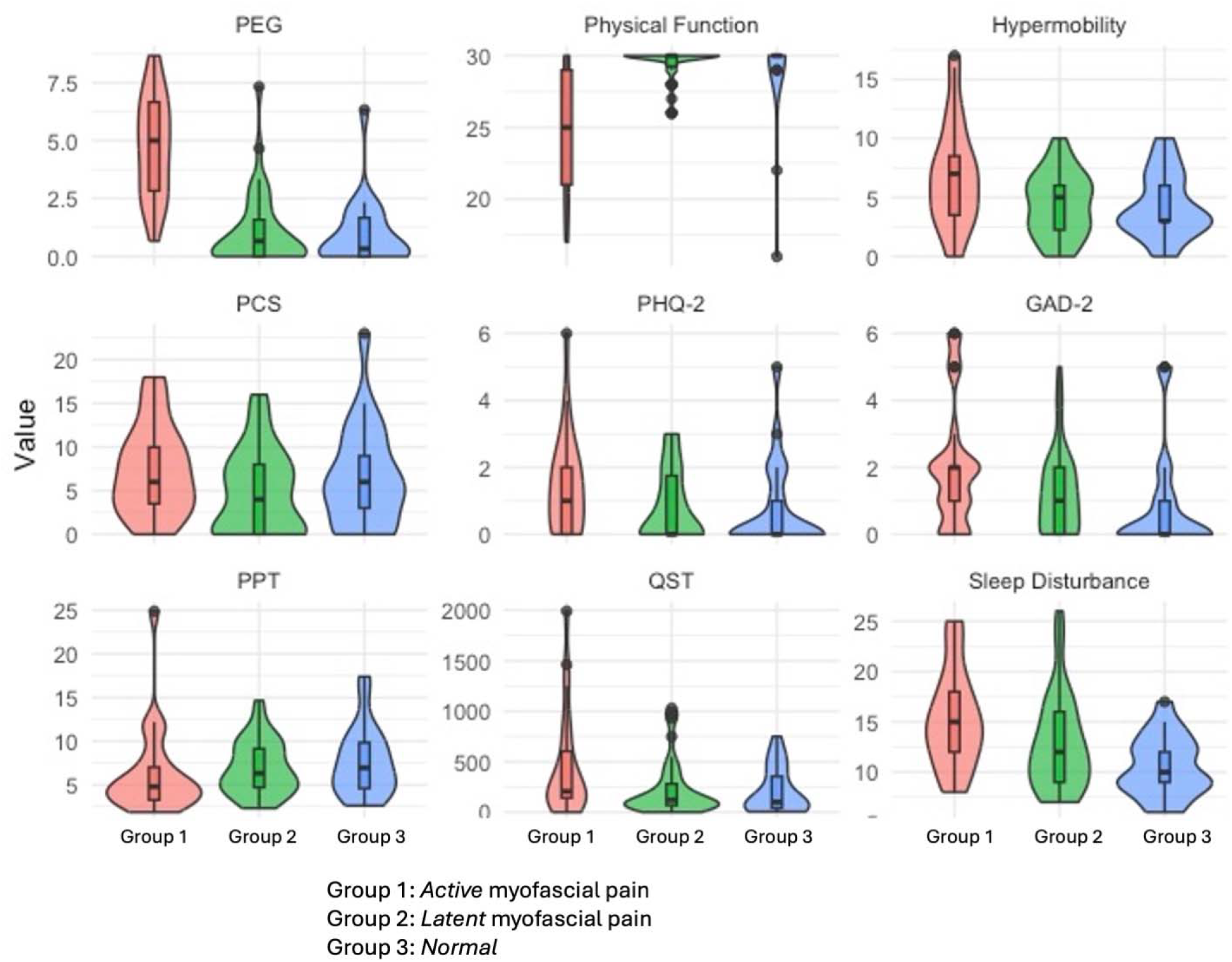
Distribution of values of patient reported outcomes and objective measures between subjects clinically classified into *active, latent* and *normal* groups. There is strong overlap on self-reported outcomes between groups, highlighting that whole-person biopsychosocial measures are not explained by regional findings on physical examination.

To explore the relationships among variables, we applied graphical LASSO to construct partial correlation networks^39,40^. The analysis was performed using the *qgraph* package in R (version 4.3.3). For the penalty term λ, we first determined the upper bound as the smallest value of λ that results in a network with no connected edges. The minimum value of λ was set to 0.01 times the upper bound, ensuring a fully connected and sufficiently complex network. A sequence of equally-spaced λ values was then generated within this range, and the optimal λ was selected by minimizing the Extended Bayesian Information Criterion (eBIC)^25^.

## Results

Out of 137 subjects screened, 96 subjects met eligibility criteria and were recruited for the study. Complete records for all variables were available for 82 subjects. Reasons for the 14 missing records were the following: (1) Did not complete multiple questionnaires (N= 5); (2) Did not answer all fields on one questionnaire (N=5; PCS: 2 incomplete records; PEG: 2 incomplete records; PHQ-2: one incomplete record); (3) Missing age (N=2); (4) Missing PPT (N=2). Subjects with complete data records for all selected variables (N=82) were included in this analysis.

### Demographic and Clinical Data

Descriptive characteristics (mean ± standard deviation or percentage) of the variables (physical findings and self-reports) in the study sample are shown in Table 1, separately for the *active, latent* and *normal* groups and for all subjects.

Current bilateral neck/shoulder pain was reported by 41.25% of the participants, while 17.5% reported unilateral pain and 41.25% of participants did not report experiencing pain currently. In our sample, 36.4% reported both neck and shoulder pain, 14.3% reported neck pain only, 11.7% reported shoulder pain only, while 36.6% reported neither neck nor shoulder pain. Physical findings of MTrPs and taut bands were ubiquitous in the sample: bilateral findings were present in 79.3% of the subjects, unilateral findings were present in 11%, and there were no findings in only 9.7%. Based on the physical exam, 28.1% were determined to have active myofascial pain, 46.3% were determined to have *latent* myofascial pain, and the remaining 25.6% had no myofascial pain. Physical findings in *normal* subjects were not considered to meet Travell and Simon’s criteria for myofascial involvement.

Individual measures exhibit large variability and overlap among subjects in the *active, latent* and *normal* groups (Figure 2). In particular, there is strong overlap on PEG between the *latent* myofascial pain and *normal* groups, demonstrating that subjects determined to be asymptomatic by myofascial criteria may have symptom burden similar to those presenting with latent MP. Based on self-reported outcomes, the *normal* group was not truly asymptomatic.

### Regression Analysis

Table 2 shows the regression analysis for PEG as a dependent variable. In the pooled sample, 75% of the variance is explained by the predictors and adjustment variables. Partial R^2^ values represent the proportion of variance explained by each predictor after adjusting for other variables. Physical Function and PCS are the most important predictors in the model, with partial R^2^ of 25.6% (p<0.0001) and 16.7% (p=0.0004), respectively. As expected, the adjustment variable, pain group, is important, so we examine the subgroup analysis defined by the group (Tables 3, 4 and 5). The predictors and adjustment variables explain 85% and 92% of the variance in the *active* and *normal* groups but only 46% of the variance in the *latent* group. In the active group, the most important predictor is PCS, with a partial R^2^ of 37.5% (p<0.02). In the normal group, Physical Function (70.7%, p=0.0006), PCS (41.9%, p=0.023), Hypermobility (35.6%, p<0.05), and GAD-2 (34.4%, p<0.05) all show substantial partial R^2^ values. In the latent group, PHQ-2 is the most important predictor, with a partial R^2^ of 20.2% (p=0.014). Sex at birth is an important adjustment variable in both the *active* and *normal groups*, but not in the latent group. Variance inflation factors indicated no evidence of substantial multicollinearity across the three regression models.

**Table 2.** Regression analysis for dependent variable PEG (R^2^ = 0.75)

| Variable | Estimate | S.E. | p-value | partial $R^2$ |
| --- | --- | --- | --- | --- |
| (Intercept) | 8.47 | 2.18 | 0.0002 | . |
| Pain Group (*active*) | 2.39 | 0.51 | <0.0001 | 0.2622 |
| Pain Group (*latent*) | 0.43 | 0.38 | 0.2565 | 0.2622 |
| Physical Function | -0.29 | 0.06 | <0.0001 | 0.2555 |
| PCS | 0.12 | 0.03 | 0.0004 | 0.1671 |
| PHQ-2 | 0.27 | 0.16 | 0.0912 | 0.0408 |
| Hypermobility | -0.06 | 0.05 | 0.2762 | 0.0172 |
| GAD-2 | -0.13 | 0.13 | 0.3408 | 0.0132 |
| Sleep Disturbance | 0.03 | 0.04 | 0.3618 | 0.0121 |
| Sex at Birth (Male) | 0.30 | 0.36 | 0.4042 | 0.0101 |
| PPT | -0.02 | 0.05 | 0.6301 | 0.0034 |
| QST | -0.0001 | 0.0005 | 0.7670 | 0.0013 |

**Table 3.** Regression analysis for dependent variable PEG in the *active myofascial pain* group (R^2^ = 0.85)

| Variable | Estimate | S.E. | p-value | partial $R^2$ |
| --- | --- | --- | --- | --- |
| (Intercept) | 8.67 | 4.07 | 0.0546 | . |
| PCS | 0.29 | 0.11 | 0.0199 | 0.3753 |
| Sex at Birth (Male) | -2.20 | 0.87 | 0.0268 | 0.3464 |
| GAD-2 | -0.50 | 0.26 | 0.0766 | 0.2383 |
| PHQ-2 | 0.37 | 0.25 | 0.1766 | 0.1466 |
| Physical Function | -0.12 | 0.10 | 0.2397 | 0.1131 |
| QST | -0.0007 | 0.0007 | 0.3168 | 0.0833 |
| Sleep Disturbance | -0.07 | 0.08 | 0.4016 | 0.0593 |
| Hypermobility | -0.07 | 0.09 | 0.4801 | 0.0424 |
| Age | -0.01 | 0.03 | 0.7483 | 0.0089 |
| PPT | 0.02 | 0.06 | 0.7521 | 0.0086 |

**Table 4.** Regression analysis for dependent variable PEG in the *latent myofascial pain* group (R^2^ = 0.46)

| Variable | Estimate | S.E. | p-value | partial $R^2$ |
| --- | --- | --- | --- | --- |
| (Intercept) | 0.41 | 6.68 | 0.9513 | . |
| PHQ-2 | 0.76 | 0.29 | 0.0143 | 0.2024 |
| PCS | 0.10 | 0.06 | 0.0862 | 0.1051 |
| Age | 0.03 | 0.02 | 0.1204 | 0.0870 |
| Sex at Birth (Male) | 0.87 | 0.59 | 0.1483 | 0.0758 |
| Hypermobility | -0.09 | 0.09 | 0.3575 | 0.0314 |
| Sleep Disturbance | 0.04 | 0.05 | 0.4651 | 0.0199 |
| GAD-2 | -0.10 | 0.23 | 0.6574 | 0.0074 |
| Physical Function | -0.06 | 0.21 | 0.7632 | 0.0034 |
| QST | -0.0002 | 0.0009 | 0.8150 | 0.0021 |
| PPT | 0.01 | 0.09 | 0.9473 | 0.0002 |

**Table 5.** Regression analysis for dependent variable PEG in the *normal* group (R^2^ = 0.92)

| Variable | Estimate | S.E. | p-value | partial $R^2$ |
| --- | --- | --- | --- | --- |
| (Intercept) | 12.08 | 3.15 | 0.0033 | . |
| Physical Function | -0.41 | 0.08 | 0.0006 | 0.7072 |
| Sex at Birth (Male) | 1.38 | 0.35 | 0.0028 | 0.6073 |
| PCS | 0.09 | 0.03 | 0.0230 | 0.4187 |
| Age | -0.05 | 0.02 | 0.0339 | 0.3762 |
| Hypermobility | 0.17 | 0.07 | 0.0405 | 0.3563 |
| GAD-2 | -0.35 | 0.15 | 0.0451 | 0.3439 |
| Sleep Disturbance | 0.06 | 0.07 | 0.4090 | 0.0691 |
| PPT | -0.02 | 0.04 | 0.6412 | 0.0226 |
| QST | 0.0003 | 0.0009 | 0.7624 | 0.0096 |
| PHQ-2 | -0.03 | 0.15 | 0.8303 | 0.0048 |

### Network Analysis

For the *active* and *latent* groups, pairwise partial correlations with graphical LASSO resulted in models with a median of 14 (interquartile range 12-15) and 4 (interquartile range 3-6) non-zero edges, respectively (Figure 3). No non-zero edges could be reliably found in the normal group. In Figure 3, for the *active* group, physical function is negatively correlated with hypermobility (−0.43) and PEG (−0.37). PCS is positively correlated with both PEG (0.38) and GAD-2 (0.36). In contrast the *latent* group exhibits a different network topology, with a negative correlation between Physical Function and PCS (−0.26), which is weakly correlated with PEG (0.06). PHQ-2 and GAD-2 remain positively correlated (0.31), while the negative correlation between PPT and QST is weak (−0.07). See Supplement for additional details.

**Figure 3.**
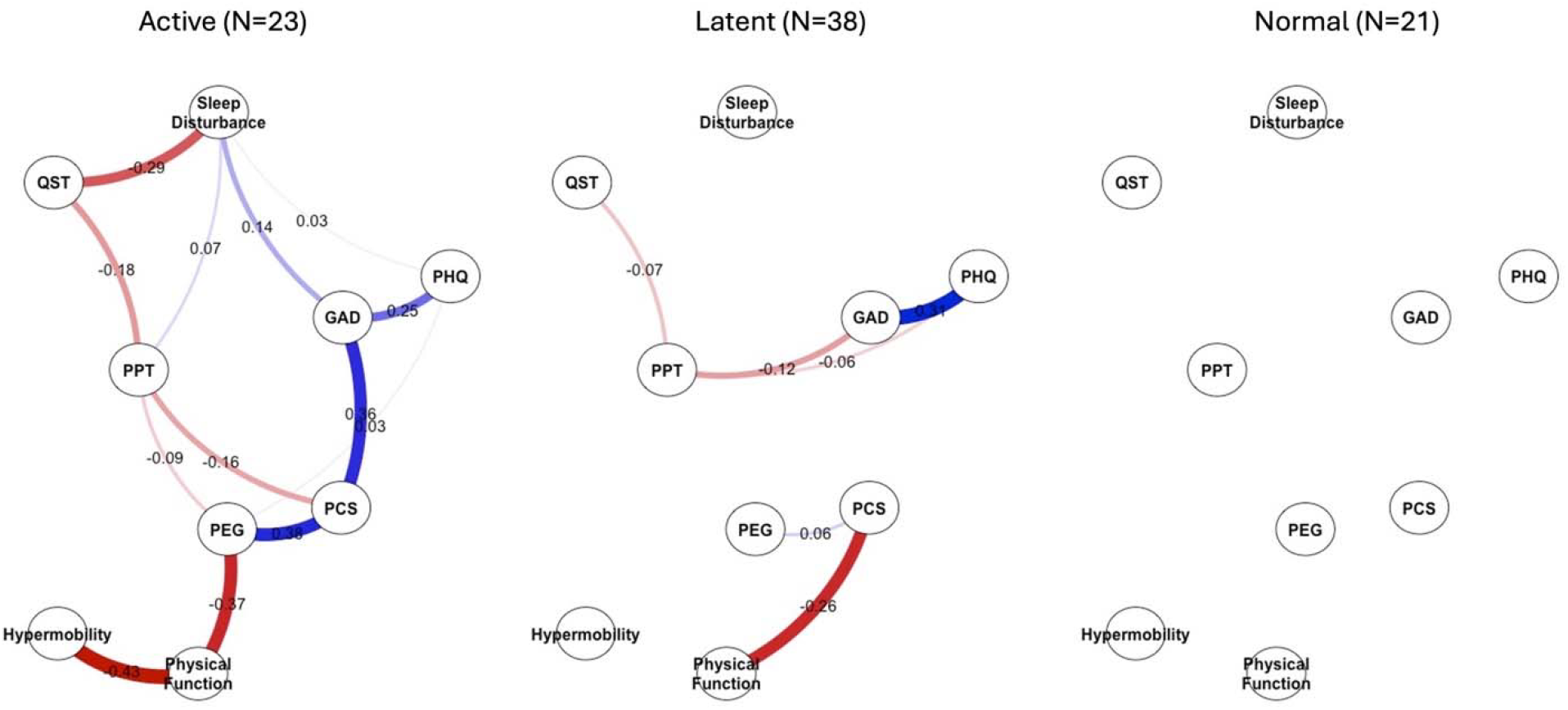
Partial correlation networks for the active, *latent* and *normal* groups. The optimal λ was selected by minimizing the standard Bayesian Information Criterion. Using jackknife methods, a median of 14 edges were found in the *active* group, and 4 edges in the *latent* group. No edges were found in the *normal* group.

## Discussion

This study presents a clinically relevant, multidimensional approach to evaluating the burden of symptoms associated with MP, in the absence of a clear consensus on how MP should be assessed or managed^8,17^.

### Key Findings and Interpretation

Our findings build upon previous literature in three important ways. First, our data revealed significant variability in the self-reports and objective findings across clinically-relevant subgroups in a cohort of subjects recruited from the general community. Second, while the variability was not unexpected since our eligibility criteria were intentionally broad, we found that a multidimensional analysis using assessments across physical and psychosocial domains explained much of the variability in pain intensity and intereference. Third, a network analysis identified linkages among clinically relevant variables (nodes) explaining potential pathways contributing to identifying which symptoms are prevalent and their variability. The active group demonstrated a high symptom burden with links between higher PEG, poorer physical function, hypermobility, higher levels of pain catastrophizing (PCS), and higher levels of anxiety and depression (GAD and PHQ-2); and higher levels of sensitization based on PPT and QST, linked to sleep disturbance. This is not the case for the latent group, which had fewer links (PHQ-GAD; and PCS-PF). In *normal* subjects, the graphical LASSO yielded a fully sparse precision matrix, with no edges retained at the selected regularization level with insufficient evidence for strong conditional associations among the variables. These collective findings provide novel early evidence supporting the need to understand how physical, emotional, social, and environmental factors all shape how symptom burden is individually experienced, and the need for prioritizing targeted interventions to improve whole-person outcomes.

Our findings in MP are consistent with previous literature on chronic pain networks^18–22^. Pain catastrophizing^41^ was shown to strengthen the association between pain and both anxiety and depression^18^. The limbic system mediates pain by integrating emotional and motivational dimensions with inputs from the somatosensory cortex^42–45^ and interacts bidirectionally with the hypothalamus-pituitary-adrenal (HPA) axis^46^. Thus, chronic stress may lead to HPA dysregulation and pain sensitivity creating a cycle of chronic pain and stress^47^. Notably, patients with chronic upper trapezius MP demonstrate increased limbic system (i.e., anterior insula) activity^45^. Our results do not imply causal links between physical and psychosocial components and the psychosocial burden may result from the stress of navigating the healthcare system when faced with the need for managing multiple symptoms^48^, which in turn can perpetuate pain catastrophizing.

One novel finding in the network structure is the link between hypermobility and physical function, which shows a strong partial correlation in the *active* group but not in the *latent* group. A higher prevalence of hypermobility in MP has been reported^49^, and our results indicate that physical function mediates the association between hypermobility and pain.

While active and latent MTrPs are currently used for classifying the physical findings associated with MP, evidence supports the hypothesis that the regional findings in MP are linked to multivariate central mechanisms that may be common across many somatic, neurologic, and visceral pain conditions^12,45^. The “myofascial unit”, defined as an integrated anatomical and functional structure that includes muscle fibers, fascia (endomysium, perimysium and epimysium) and its associated innervations (free nerve endings, muscle spindles), lymphatics, and blood vessels^23,24,50,51^, has been implicated to play a foundational role^8,9,12,52–61^.

Experimental data help link mechanisms of central sensitization, a common component of all chronic pain syndromes, to dysfunction at the myofascial unit through the mechanism of neurogenic inflammation^58,59,62,63^. For example, several studies have reported soft tissue findings associated with pain conditions of visceral hypersensitivity such as irritable bowel syndrome and chronic pelvic pain^64–66^, which raises the possibility that a primary pathology residing within somatic or visceral tissues^59^, such as facet joints, irritable bowel, and endometriosis, etc. could contribute to dysfunction of the myofascial unit.

### Implications for Future Research

A critical impediment to clinical and mechanistic research in MP is the lack of reliable and reproducible diagnostic criteria^10,11,13,17^. While imaging and other biomarkers could improve reproducibility^54^, the clinical variables we selected for this study^26^, explained 75% of the variance of PEG in our sample of patients with MP. In contrast, the presence of physical findings was ubiquitous (Table 1). Our findings provide evidence for a shift away from relying solely on the unreliable identification of MTrPs and other regional pain contributors towards assessments that might provide insight into managing the symptom burden of the whole person. Future research needs to investigate individual networks of causal linkages from longitudinal observations of relevant biopsychosocial variables to understand individual differences in pain perception across time^67,68^, including in subclinical latent phenotypes, and may lower the number of patients labeled non-specific cMSKP^3^. The links between the central and peripheral nervous system and the myofascial unit need further exploration.

### Implications for Clinical Management

Traditional clinical practice in MP has been siloed by regional symptomatology, with an emphasis on MTrPs as the target for assessment and treatment. This approach fails to adequately capture the complex reciprocal systemic biopsychosocial interactions, making it challenging to identify which treatments are likely to work for which subject. While psychosocial symptoms are indicative of poor outcomes^69–74^, current predictive and prognostic models^75,76^ do not provide insight into how to alter outcomes.

Network models might provide a new approach to meaningfully integrate information across biopsychosocial domains to guide mechanism-informed clinical decision-making^19,20^. Key central influencers within individual networks can offer targets for multidisciplinary therapeutic interventions^77,78^. It is our view that the larger symptom burden of the active clinical profile is likely to require earlier, and more intense and comprehensive care and may experience a longer recovery time compared to the latent profile which exhibited lower symptom burden. Identifying patients with higher risk profiles can avoid unnecessary testing and interventions for lower risk individuals.

### Limitations

Potential sources of bias include incomplete records from 15% of recruited subjects, and more women in the *active* group. While the overall sample size is adequate for regression, the sample size of the *active* group was small for a multivariate network analysis. These findings need to be confirmed in a larger sample. The network stability was moderate for this sample size although the network structure was not sensitive to parameter choice (see Supplementary material). Our analyses were exploratory without adjustment for multiple testing, so p-values should serve as a guide rather than a formal level of significance.

## Conclusion

Self-reports and objective measures highlight a complex biopsychosocial burden of symptoms in the patient with MP and explains the observed variability in pain intensity and interference. Network analysis can elucidate interconnections and mediation among multidimensional factors and provides an innovative whole-person clinical approach for risk stratification driven by symptom burden that does not solely rely on identifying specific local pain contributors. Our findings can inform future longitudinal studies aimed at elucidating the directionality, temporality and predictive value of biopsychosocial contributors to the clinical spectrum of chronic non-specific cMSKP with myofascial involvement.

## Data Availability

All data produced in the present study are available upon reasonable request to the authors

## Abbreviations

(MP): Myofascial Pain
(cMSKP): chronic musculoskeletal pain
(MTrP): myofascial trigger point
(PEG): Pain intensity and interference
(PCS): Pain catastrophizing
(PF): PROMIS physical function
(GAD-2): generalized anxiety disorder
(PHQ-2): Patient health questionnaire
(PPT): Pressure pain threshold
(QST): Quantitative Sensory Testing
(HEAL): Helping End Addiction Long Term
(CDE): Common Data Elements

## SUPPLIERS

1. Pressure algometer (Commander Algometer, JTech Medical, Salt Lake City, UT)
2. The PinPrick Stimulator Set (MRC Systems, Heidelberg, Germany)

## Notes

Conflicts of Interest: All sources of funding and other relationships disclosed in attached ICJME form

### Competing Interest Statement

This work was supported by NIH R61AT012286 Multimodal imaging biomarkers for investigating fascia, muscle and vasculature in myofascial pain.
María José Guzmán Pavón received support from the University of Castilla-La Mancha to travel to George Mason University to participate in this research.
Secili DeStefano has received consulting fees from George Mason University as part of the NIH grant NIH R61AT012286 to participate in this research.
Siddhartha Sikdar, Lynn Gerber and Secili DeStefano are members of the George Mason University Chronic Pain Community Advisory Board (CPAB). Patient advocates, who are members of the CPAB, were involved as research partners in helping refine the research question, understanding the gap between lived experience of chronic pain and academic research, emphasizing the need for appropriate nomenclature, and reviewing the study findings.
Siddhartha Sikdar, Lynn Gerber, Jay Shah, Antonio Stecco, John Srbely and Secili DeStefano are members of the ISPRM PAIN Special Interest Group

### Clinical Protocols

https://clinicaltrials.gov/study/NCT06060925

### Funding Statement

This work was supported by NIH R61AT012286 and R01 AT013603

### Author Declarations

Institutional Review Board of George Mason University gave ethical approval for this work

## REFERENCES

1. Ferreira, M. L. et al. Global, regional, and national burden of low back pain, 1990–2020, its attributable risk factors, and projections to 2050: a systematic analysis of the Global Burden of Disease Study 2021. Lancet Rheumatol. 5, e316–e329 (2023).

2. Steinmetz, J. D. et al. Global, regional, and national burden of osteoarthritis, 1990–2020 and projections to 2050: a systematic analysis for the Global Burden of Disease Study 2021. Lancet Rheumatol. 5, e508–e522 (2023).

3. Balagué, F., Mannion, A. F., Pellisé, F. & Cedraschi, C. Non-specific low back pain. The Lancet 379, 482–491 (2012).

4. Dor, A. & Kalichman, L. A myofascial component of pain in knee osteoarthritis. J. Bodyw. Mov. Ther. 21, 642–647 (2017).

5. Iglesias-González, J. J., Muñoz-García, M. T., Rodrigues-de-Souza, D. P., Alburquerque-Sendín, F. & Fernández-de-las-Peñas, C. Myofascial Trigger Points, Pain, Disability, and Sleep Quality in Patients with Chronic Nonspecific Low Back Pain. Pain Med. 14, 1964–1970 (2013).

6. Ezzati, K. et al. Prevalence of Cervical Myofascial Pain Syndrome and its Correlation with the Severity of Pain and Disability in Patients with Chronic Non-specific Neck Pain. Arch. Bone Jt. Surg. 9, 230– 234 (2021).

7. Gerwin, R. D. Classification, epidemiology, and natural history of myofascial pain syndrome. Curr. Pain Headache Rep. 5, 412–420 (2001).

8. Shah, J. P. et al. Myofascial Trigger Points Then and Now: A Historical and Scientific Perspective. PM R 7, 746–761 (2015).

9. Gerber, L. H. et al. A systematic comparison between subjects with no pain and pain associated with active myofascial trigger points. PM R 5, 931–938 (2013).

10. Hsieh, C.-Y. J. et al. Interexaminer reliability of the palpation of trigger points in the trunk and lower limb muscles. Arch. Phys. Med. Rehabil. 81, 258–264 (2000).

11. Myburgh, C., Larsen, A. H. & Hartvigsen, J. A Systematic, Critical Review of Manual Palpation for Identifying Myofascial Trigger Points: Evidence and Clinical Significance. Arch. Phys. Med. Rehabil. 89, 1169–1176 (2008).

12. Sikdar, S. et al. A model for personalized diagnostics for non-specific low back pain: the role of the myofascial unit. Front. Pain Res. 4, (2023).

13. Rivers, W. E., Garrigues, D., Graciosa, J. & Harden, R. N. Signs and Symptoms of Myofascial Pain: An International Survey of Pain Management Providers and Proposed Preliminary Set of Diagnostic Criteria. Pain Med. Malden Mass 16, 1794–1805 (2015).

14. Park, S. J., Yoon, D. M., Yoon, K. B., Moon, J. A. & Kim, S. H. Factors Associated with Higher Reported Pain Levels in Patients with Chronic Musculoskeletal Pain: A Cross-Sectional, Correlational Analysis. PLOS ONE 11, e0163132 (2016).

15. Finestone, H. M., Alfeeli, A. & Fisher, W. A. Stress-induced Physiologic Changes as a Basis for the Biopsychosocial Model of Chronic Musculoskeletal Pain: A New Theory? Clin. J. Pain 24, 767 (2008).

16. Armstrong, M., Castellanos, J. & Christie, D. Chronic pain as an emergent property of a complex system and the potential roles of psychedelic therapies. Front. Pain Res. 5, (2024).

17. Phan, V. et al. Myofascial Pain Syndrome: A Narrative Review Identifying Inconsistencies in Nomenclature. PM R 12, 916–925 (2020).

18. Wi, D., Park, C., Ransom, J. C., Flynn, D. M. & Doorenbos, A. Z. A network analysis of pain intensity and pain-related measures of physical, emotional, and social functioning in US military service members with chronic pain. Pain Med. Malden Mass 25, 231–238 (2024).

19. Zhou, Y. et al. Network Analysis of Pain Catastrophizing, Self-Efficacy, and Kinesiophobia Among Patients After Total Knee Arthroplasty: A Cross-Sectional Study. Patient Prefer. Adherence 18, 1897– 1906 (2024).

20. Solmi, M. et al. Network analysis of the relationship between depressive symptoms, demographics, nutrition, quality of life and medical condition factors in the Osteoarthritis Initiative database cohort of elderly North-American adults with or at risk for osteoarthritis. Epidemiol. Psychiatr. Sci. 29, e14 (2020).

21. Gevers-Montoro, C. et al. A network analysis on biopsychosocial factors and pain-related outcomes assessed during a COVID-19 lockdown. Sci. Rep. 13, 4399 (2023).

22. Valera-Calero, J. A., Arendt-Nielsen, L., Cigarán-Méndez, M., Fernández-de-las-Peñas, C. & Varol, U. Network Analysis for Better Understanding the Complex Psycho-Biological Mechanisms behind Fibromyalgia Syndrome. Diagnostics 12, 1845 (2022).

23. Stecco, A. et al. From Muscle to the Myofascial Unit: Current Evidence and Future Perspectives. Int. J. Mol. Sci. 24, 4527 (2023).

24. Langevin, H. M. Fascia Mobility, Proprioception, and Myofascial Pain. Life 11, 668 (2021).

25. Borsboom, D. et al. Network analysis of multivariate data in psychological science. Nat. Rev. Methods Primer 1, 1–18 (2021).

26. Adams, M. C. B. et al. NIH HEAL Common Data Elements (CDE) implementation: NIH HEAL Initiative IDEA-CC. Pain Med. 24, 743–749 (2023).

27. Travell, J. G. & Simons, D. G. Myofascial Pain and Dysfunction: The Trigger Point Manual. (Lippincott Williams & Wilkins, 1983).

28. Krebs, E. E. et al. Development and Initial Validation of the PEG, a Three-item Scale Assessing Pain Intensity and Interference. J. Gen. Intern. Med. 24, 733–738 (2009).

29. Sullivan, M. J. L., Bishop, S. R. & Pivik, J. The Pain Catastrophizing Scale: Development and validation. Psychol. Assess. 7, 524–532 (1995).

30. Yee, T. J. et al. Correlation between the Oswestry Disability Index and the 4-item short forms for physical function and pain interference from PROMIS. (2019) doi:10.3171/2019.5.SPINE19400.

31. Buysse, D. J. et al. Development and validation of patient-reported outcome measures for sleep disturbance and sleep-related impairments. Sleep 33, 781–792 (2010).

32. Bisby, M. A. et al. Examining the psychometric properties of brief screening measures of depression and anxiety in chronic pain: The Patient Health Questionnaire 2-item and Generalized Anxiety Disorder 2-item. Pain Pract. Off. J. World Inst. Pain 22, 478–486 (2022).

33. Bockhorn, L. N. et al. Interrater and Intrarater Reliability of the Beighton Score: A Systematic Review. Orthop. J. Sports Med. 9, 2325967120968099 (2021).

34. Beighton, P. & Horan, F. Orthopaedic aspects of the Ehlers-Danlos syndrome. J. Bone Joint Surg. Br. 51, 444–453 (1969).

35. Juul-Kristensen, B., Røgind, H., Jensen, D. V. & Remvig, L. Inter-examiner reproducibility of tests and criteria for generalized joint hypermobility and benign joint hypermobility syndrome. Rheumatology 46, 1835–1841 (2007).

36. Park, G., Kim, C. W., Park, S. B., Kim, M. J. & Jang, S. H. Reliability and usefulness of the pressure pain threshold measurement in patients with myofascial pain. Ann. Rehabil. Med. 35, 412–417 (2011).

37. Cheatham, S. W., Kolber, M. J., Mokha, G. M. & Hanney, W. J. Concurrent validation of a pressure pain threshold scale for individuals with myofascial pain syndrome and fibromyalgia. J. Man. Manip. Ther. 26, 25–35 (2018).

38. Allison, C., Korey, L. & John Z S., A novel computational technique for the quantification of temporal summation in healthy individuals. Musculoskelet. Sci. Pract. 54, 102400 (2021).

39. Foygel, R. & Drton, M. Extended Bayesian Information Criteria for Gaussian Graphical Models. in Advances in Neural Information Processing Systems vol. 23 (Curran Associates, Inc., 2010).

40. Epskamp, S. & Fried, E. I. A tutorial on regularized partial correlation networks. Psychol. Methods 23, 617–634 (2018).

41. Petrini, L. & Arendt-Nielsen, L. Understanding Pain Catastrophizing: Putting Pieces Together. Front. Psychol. 11, (2020).

42. Meade, E. & Garvey, M. The Role of Neuro-Immune Interaction in Chronic Pain Conditions; Functional Somatic Syndrome, Neurogenic Inflammation, and Peripheral Neuropathy. Int. J. Mol. Sci. 23, 8574 (2022).

43. Frischenschlager, O. & Pucher, I. Psychological management of pain. Disabil. Rehabil. 24, 416–422 (2002).

44. Aronoff, G. & Feldman, J. Preventing disability from chronic pain: a review and reappraisal. Int. Rev. Psychiatry 12, 157–169 (2000).

45. Niddam, D. M., Chan, R.-C., Lee, S.-H.Yeh, T.-C. & Hsieh, J.-C. Central representation of hyperalgesia from myofascial trigger point. NeuroImage 39, 1299–1306 (2008).

46. Herman, J., Cullinan, W. & Prewitt, C. Neuronal circuit regulation of the hypothalamo-pituitary-adrenocortical stress axis. Crit. Rev. Neurobiol. 10, 371–394 (1996).

47. Wyns, A., Nijs, J. & Godderis, L. The Biology of Stress Intolerance in Patients with Chronic Pain-State of the Art and Future Directions. J. Clin. Med. 12, 2245 (2023).

48. Costa, N. et al. “I felt uncertain about my whole future”—a qualitative investigation of people’s experiences of navigating uncertainty when seeking care for their low back pain. PAIN 164, 2749 (2023).

49. Tuna, M. The Evaluation of the Frequency of Benign Joint Hypermobility in Patients with Myofascial Pain Syndrome. https://www.turkosteoporozdergisi.org/articles/miyofasiyal-agri-sendromlu-hastalarda-eklem-hipermobilitesi-sikliginin-degerlendirilmesi/doi/tod.galenos.2022.80958 (2023) doi:10.4274/tod.galenos.2022.80958.

50. Tuckey, B., Srbely, J., Rigney, G., Vythilingam, M. & Shah, J. Impaired Lymphatic Drainage and Interstitial Inflammatory Stasis in Chronic Musculoskeletal and Idiopathic Pain Syndromes: Exploring a Novel Mechanism. Front. Pain Res. 2, 45 (2021).

51. Langevin, H. M. et al. Reduced thoracolumbar fascia shear strain in human chronic low back pain. BMC Musculoskelet. Disord. 12, 203 (2011).

52. Shah, J. P., Phillips, T. M., Danoff, J. V. & Gerber, L. H. An in vivo microanalytical technique for measuring the local biochemical milieu of human skeletal muscle. J. Appl. Physiol. 99, 1977–1984 (2005).

53. Shah, J. P. et al. Biochemicals Associated With Pain and Inflammation are Elevated in Sites Near to and Remote From Active Myofascial Trigger Points. Arch. Phys. Med. Rehabil. 89, 16–23 (2008).

54. Sikdar, S. et al. Novel Applications of Ultrasound Technology to Visualize and Characterize Myofascial Trigger Points and Surrounding Soft Tissue. Arch. Phys. Med. Rehabil. 90, 1829–1838 (2009).

55. Sikdar, S., Ortiz, R., Gebreab, T., Gerber, L. H. & Shah, J. P. Understanding the vascular environment of myofascial trigger points using ultrasonic imaging and computational modeling. Annu. Int. Conf. IEEE Eng. Med. Biol. Soc. IEEE Eng. Med. Biol. Soc. Annu. Int. Conf. 2010, 5302–5305 (2010).

56. Turo, D. et al. Novel Use of Ultrasound Elastography to Quantify Muscle Tissue Changes After Dry Needling of Myofascial Trigger Points in Patients With Chronic Myofascial Pain. J Ultrasound Med 34, 2149–2161 (2015).

57. Gerber, L. H. et al. Dry Needling Alters Trigger Points in the Upper Trapezius Muscle and Reduces Pain in Subjects With Chronic Myofascial Pain. PM&R 7, 711–718 (2015).

58. Srbely, J. Z., Dickey, J. P. & Montaholi, Y. Experimentally Induced Central Sensitization Evokes Segmental Autonomic Responses in Humans. Int. Phys. Med. Rehabil. J. 1, 1–7 (2017).

59. Srbely, J. Z., Dickey, J. P., Bent, L. R., Lee, D. & Lowerison, M. Capsaicin-Induced Central Sensitization Evokes Segmental Increases in Trigger Point Sensitivity in Humans. J. Pain 11, 636–643 (2010).

60. Srbely, J., Dickey, J., Lee, D. & Lowerison, M. Dry needle stimulation of myofascial trigger points evokes segmental anti-nociceptive effects. J. Rehabil. Med. 42, 463–468 (2010).

61. Gerber, L. H. et al. Beneficial Effects of Dry Needling for Treatment of Chronic Myofascial Pain Persist for 6 Weeks After Treatment Completion. PM R 9, 105–112 (2017).

62. Duarte, F. C. K. et al. Experimentally induced spine osteoarthritis in rats leads to neurogenic inflammation within neurosegmentally linked myotomes. Exp. Gerontol. 149, 111311 (2021).

63. Duarte, F. C. K., Hurtig, M., Clark, A., Simpson, J. & Srbely, J. Z. Association between naturally occurring spine osteoarthritis in geriatric rats and neurogenic inflammation within neurosegmentally linked skeletal muscle. Exp. Gerontol. 118, 31–38 (2019).

64. Gerwin, R. D. Myofascial and Visceral Pain Syndromes: Visceral-Somatic Pain Representations. J. Musculoskelet. Pain 10, 165–175 (2002).

65. Doggweiler-Wiygul, R. Urologic myofascial pain syndromes. Curr. Pain Headache Rep. 8, 445–451 (2004).

66. Stratton, P., Khachikyan, I., Sinaii, N., Ortiz, R. & Shah, J. Association of Chronic Pelvic Pain and Endometriosis With Signs of Sensitization and Myofascial Pain. Obstet. Gynecol. 125, 719–728 (2015).

67. Von Korff, M., Dworkin, S. F., Le Resche, L. & Kruger, A. An epidemiologic comparison of pain complaints. PAIN 32, 173 (1988).

68. Affleck, G., Tennen, H., Urrows, S. & Higgins, P. Individual differences in the day-to-day experience of chronic pain: A prospective daily study of rheumatoid arthritis patients. Health Psychol. 10, 419– 426 (1991).

69. Go, D. J. et al. Metabolic obesity and the risk of knee osteoarthritis progression in elderly community residents: A 3-year longitudinal cohort study. Int. J. Rheum. Dis. 25, 192–200 (2022).

70. Hall, A. J., Stubbs, B., Mamas, M. A., Myint, P. K. & Smith, T. O. Association between osteoarthritis and cardiovascular disease: Systematic review and meta-analysis. Eur. J. Prev. Cardiol. 23, 938–946 (2016).

71. Aqeel, M., Rehna, T. & Sarfraz, R. The association among perception of osteoarthritis with adverse pain anxiety, symptoms of depression, positive and negative affects in patients with knee osteoarthritis: a cross sectional study. J. Pak. Med. Assoc. 71, 645–650 (2021).

72. Poon, C. L.-L. et al. Associations of the modified STarT back tool and Hospital Anxiety and Depression Scale (HADS) with gait speed and knee pain in knee osteoarthritis: a retrospective cohort study. Disabil. Rehabil. 44, 4452–4458 (2022).

73. Reyes, C. et al. Association Between Overweight and Obesity and Risk of Clinically Diagnosed Knee, Hip, and Hand Osteoarthritis: A Population-Based Cohort Study. Arthritis Rheumatol. Hoboken NJ 68, 1869–1875 (2016).

74. Pacca, D. M., De-Campos, G. C., Zorzi, A. R., Chaim, E. A. & De-Miranda, J. B. PREVALENCE OF JOINT PAIN AND OSTEOARTHRITIS IN OBESE BRAZILIAN POPULATION. ABCD Arq. Bras. Cir. Dig. São Paulo 31, e1344 (2018).

75. Forsythe, M. E., Dunbar, M. J., Hennigar, A. W., Sullivan, M. J. & Gross, M. Prospective Relation between Catastrophizing and Residual Pain following Knee Arthroplasty: Two-Year Follow-Up. Pain Res. Manag. 13, 730951 (2008).

76. Lewis, G. N., Rice, D. A., McNair, P. J. & Kluger, M. Predictors of persistent pain after total knee arthroplasty: a systematic review and meta-analysis.

77. Ojala, T. et al. Chronic pain affects the whole person – a phenomenological study. Disabil. Rehabil. 37, 363–371 (2015).

78. Mallick-Searle, T., Sharma, Kristin, Toal, Philip & and Gutman, A. Pain and Function in Chronic Musculoskeletal Pain—Treating the Whole Person. J. Multidiscip. Healthc. 14, 335–347 (2021).

