## Supplemental Text for "Multidimensional analysis of the clinical spectrum and symptom burden of unexplained myofascial pain"

**Supplement**

*Hyperparameter tuning and robustness of network analysis*

For the network analysis in this study, we employed the setting (penalty term $\gamma=0$), i.e., standard Bayesian information criterion. In the *active, latent* and *normal* groups, the grid search range for $\lambda$ are 0.0006-to-0.6677, 0.0006-to-0.6045 and 0.0006-to-0.6068, respectively. While there is no standard guideline for the specification of $\gamma$, the literature suggests setting $\gamma=0$ in low-dimensional settings where the number of biomarkers are smaller than the sample size while setting $\gamma=0.5$ is recommended for high-dimensional problems where there are much more biomarkers than samples^1^. The non-zero value of $\gamma$ is necessary in the high-dimensional context to reduce the number of false positives^1^. Additional tools such as EBIC, stability approach to regularization selection (StARS)^2^ k-folds cross-validation, and rotation information criterion (RIC) may be used to select the appropriate $\gamma$, but there is a lack of consensus^3^. In this study, we elected to match $\gamma$ to the data dimensionality as recommended by Foygel and Drton^1^.

We provide results from a sensitivity analysis using a non-zero penalty term ($\gamma=0.1$), which differes from the zero-penalty term used in manuscript. Applying the extended Bayesian Information Criterion (eBIC) with penalty $\gamma=0.1$imposes a stronger penalty for model complexity than the standard Bayesian Information Criterion. The resulting network for the *active* group (left panel of Figure S.2) is identical to the network in Figure 1, indicating that the partial pairwise correlations persist despite the penalty. In contrast, the network for the *currently asymptomatic* group changes with respect to the penalty. Weaker partial correlations are shrunk to zero under the eBIC criterion, resulting in a sparser network structure (middle panel of Figure S.1).

Figure S.1. An example partial correlation network of the *active*, *latent, and normal* groups obtained using the extended Bayesian Information Criterion (eBIC) with $\gamma=0.1$ to select the optimal λ. This example highlights that the weak correlations in the *latent* group can lead to a very sparse network with a certain parameter choice, but the *active* group continues to show an identical network as in Figure 1 for the default case of $\gamma=0$*.*

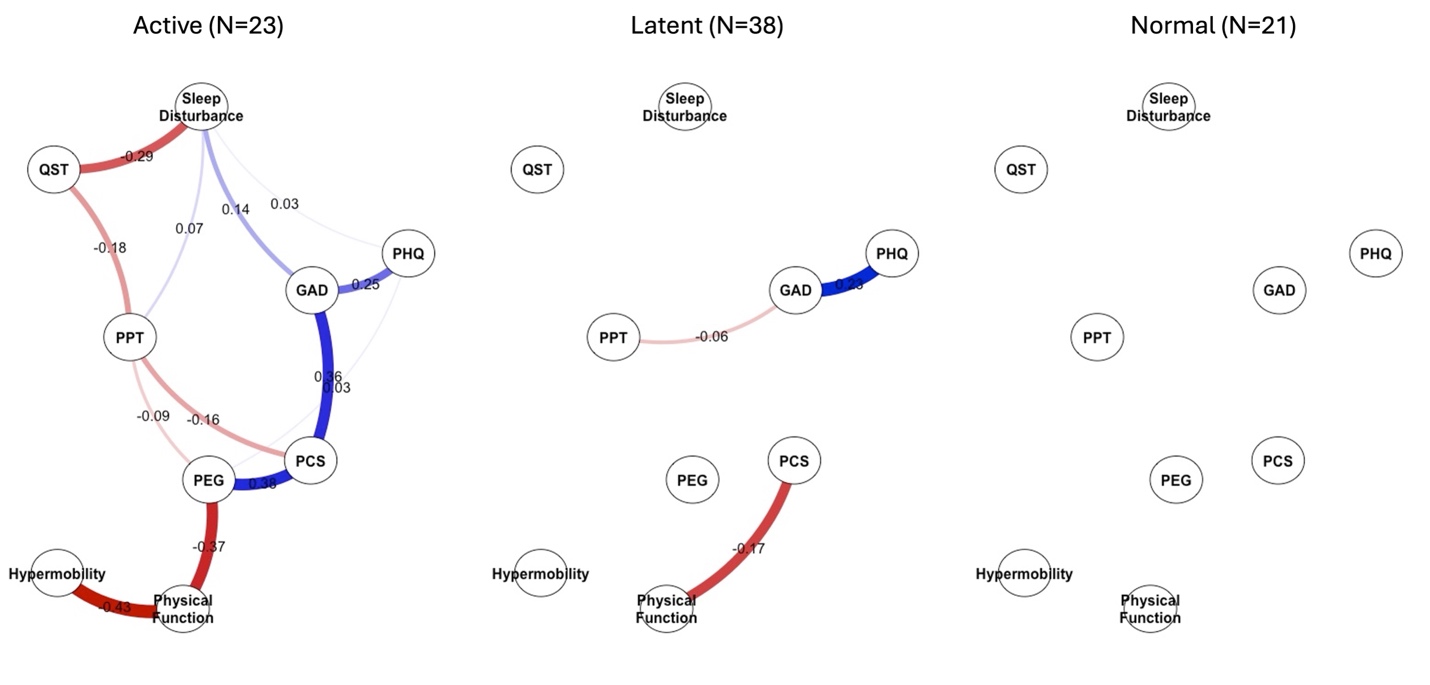

As part of our internal stability analysis, we examined the resulting network structures under different penalty terms ($\gamma$) ranging from 0-to-0.25. For each value of $\gamma$, we fit the graphical LASSO model on jack-knife samples, and we chose the optimal value of $\lambda$ based on the eBIC. For each jack-knife sample, we report the number of statistically significant edges. Table S.1 presents the median and interquartile range of the number of statistically significant edges identified across all samples for each specified $\gamma$ value. These statistics are reported separately for the active, latent and normal groups.

The chosen value $\gamma=0$ from the main manuscript provides stable results with relatively narrow interquartile ranges compared to other values. Though developing an algorithm to automatically select the penalty parameter ($\gamma$) would be a valuable contribution, it remains an open research problem and is beyond the scope of this study.

Table S.1. Median and interquartile range of the number of statistically significant edges identified across all jack-knife samples for each specified $\gamma$ value.

|  | **Active** | | | **Latent** | | | **Normal** | | |
| --- | --- | --- | --- | --- | --- | --- | --- | --- | --- |
| $\gamma$ | **Median** | **IQR** | **Width** | **Median** | **IQR** | **Width** | **Median** | **IQR** | **Width** |
| **0** | 14 | (12 , 15) | 3 | 4 | (3 , 6) | 3 | 0 | (0 , 0) | 0 |
| **0.05** | 12 | (11.5 , 13) | 1.5 | 3 | (3 , 4) | 1 | 0 | (0 , 0) | 0 |
| **0.1** | 12 | (11 , 13) | 2 | 3 | (2 , 3) | 1 | 0 | (0 , 0) | 0 |
| **0.15** | 11 | (5.5 , 12) | 6.5 | 2 | (2 , 3) | 1 | 0 | (0 , 0) | 0 |
| **0.2** | 0 | (0 , 7) | 7 | 2 | (0 , 2) | 2 | 0 | (0 , 0) | 0 |
| **0.25** | 0 | (0 , 0) | 0 | 0 | (0 , 0) | 0 | 0 | (0 , 0) | 0 |

1.	Foygel, R. & Drton, M. Extended Bayesian Information Criteria for Gaussian Graphical Models. in *Advances in Neural Information Processing Systems* vol. 23 (Curran Associates, Inc., 2010).

2.	Liu, H., Roeder, K. & Wasserman, L. Stability Approach to Regularization Selection (StARS) for High Dimensional Graphical Models. in *Advances in Neural Information Processing Systems* vol. 23 (Curran Associates, Inc., 2010).

3.	Wysocki, A. C. & Rhemtulla, M. On penalty parameter selection for estimating network models. *Multivar. Behav. Res.* **56**, 288–302 (2021).

4.	Jr, D. W. H., Lemeshow, S. & Sturdivant, R. X. *Applied Logistic Regression*. (John Wiley & Sons, 2013).
