## Appendix for "Multidimensional analysis of the clinical spectrum and symptom burden of unexplained myofascial pain"

*Details of Outcome Measures*

**Patient reported outcome measures.**

Pain, Enjoyment of Life and General activity score (PEG). An average score of three items assessing average past-week pain intensity, past-week pain interference with enjoyment of life, and past-week pain interference with general activity. This was chosen as our primary outcome because it captures both intensity and interference, the two domains our study aims to assess.

Pain Catastrophizing Scale (PCS). A 13-item questionnaire for measuring three aspects of catastrophic cognitions about pain—rumination, magnification, and helplessness. Each item was rated on a 5-point Likert scale, with a score of 4 indicating "all the time" and a score of 0 indicating "not at all."

PROMIS Physical Function. A 4-item scale to assess participants’ current self-reported capability to perform common daily activities.

PROMIS Sleep Disturbance Scale. Self-reported perceptions of sleep quality, sleep depth, and difficulties related to getting and staying asleep over a 7-day period.

Generalized Anxiety Disorder Scale 2-item scale (GAD-2). Presence of generalized anxiety based on two cognitive items scored from 0 (not at all) to 3 (nearly every day).

Patient Health Questionnaire 2 (PHQ-2). Frequency of depressed mood over the last two weeks, with a score ranging from 0 to 6.

**Modified Beighton-Brighton score**. A modified Beighton-Brighton 19-point score was performed to assess hypermobility syndrome. The 9-point Beighton score quantifies the ability to perform hyperextension tests to assess generalized joint hypermobility, while the Brighton modified criteria adds 10 additional clinical criteria for benign joint hypermobility syndrome.

**Pressure pain threshold (PPT)**. A pressure algometer (Commander Algometer, JTech Medical, Salt Lake City, UT) was used to measure minimum pressure needed to evoke discomfort on the medial upper trapezius muscle with the participant seated following standard procedures we have previously reported.

**Quantitative sensory testing (QST) for windup.** Weighted pinprick testing was performed to quantify the magnitude of temporal summation (or windup) and identify the physiological expression of excitatory post-synaptic potentials, which play an important role in central sensitization. The PinPrick Stimulator Set (MRC Systems, Heidelberg, Germany) was used to administer a series of 16 mechanical punctate stimuli at 256mN applied at 1.0 Hz perpendicular to the skin in the most sensitive area of the upper trapezius muscle bilaterally, with participants seated. Pain ratings after each pinprick were recorded on an 11-point (0–10) numeric pain rating scale.
